# T Cell Clonal Groups are Broadly Dispersed in Colon, Phenotypically Diverse, and Altered in Ulcerative Colitis

**DOI:** 10.64898/2026.04.10.26350469

**Authors:** Jeremy Fischer, Matthew P. Spindler, Graham J. Britton, Jared Weiler, Michael Tankelevich, Daniel Dai, Pablo Canales-Herrerias, Divya Jha, Urvija Rajpal, Saurabh Mehandru, Jeremiah J. Faith

## Abstract

Our understanding of human mucosal T cell clonotype distribution in health and disease has centered on immunodominant antigens. We performed single cell T cell receptor (TCR) and RNA sequencing as an untargeted approach to define distributions of T cell clonal groups in health and ulcerative colitis (UC) across 333,088 T cells in colon and peripheral blood. Healthy donor-specific TCR repertoires had limited blood-colon clonal sharing, which was highest in cytotoxic T effector memory (Tem) populations and lowest in regulatory T cells (Tregs), reflecting tissue-based compartmentalization. Within healthy colon, TCR repertoires showed high T cell clonal sharing independent of anatomic distance, associated with high intra-clonal phenotypic diversity. Colon cytotoxic and Th17 populations showed high dispersion across sites, while Tregs were compartmentalized. Clonal lineages dispersed across blood and colon upregulated trafficking markers, suggesting active movement between tissues, while those dispersed across colon sites upregulated residency markers, suggesting intra-colon repertoire sharing is mediated by long-term, slow moving clonal groups. In UC, Tregs were expanded across inflamed sites, and increased CD8 Tem clonal groups showed increased dispersion regardless of inflammation. These findings reveal principles of T cell clonal organization in the human colon during health and disease, identifying opposing patterns of clonal dispersion among Treg and Th17 clonal groups, high phenotypic diversity within dispersed clonal groups, and elevated cross-colon dispersion of CD8 Tem clonotypes in UC.

## Introduction

T cells generate antigen-specific immune responses and persistent memory through the T cell receptor (TCR). With over 10^18^ possible outcomes of variable (V), diversity (D), junction (J) gene recombination (V(D)J recombination) in αβ Τ cells (Venturi 2008), cells of the same TCR, or clonotype, are generally considered descendants of a common mature thymic emigrant. As such, multi-site single cell TCR sequencing can quantify clonal dispersion patterns across the body with each expanded clonotype representing a previous or ongoing antigen-specific response to a non-deorphaned epitope. The majority of T cells reside in lymphoid tissue, with a small fraction in blood and nonlymphoid tissues at any given time (Sender 2023). Blood T cell populations are widely studied but are generally not representative of tissue in terms of phenotype or TCR repertoire (Farber 2021, Sureshchandra 2025, Thome 2014). In addition, tissue-resident memory T cells (Trm) are a major component of mucosal immune surveillance, which are characterized by long-term survival, low mobility, and absence in circulation (Mueller 2015, Masopust 2016, Bartolomé-Casado 2019).

T cells are implicated in the pathogenesis of inflammatory bowel disease (IBD). T cells with identical antigen specificity can assume different polarization states across body sites and disease states (Sureshchandra 2025, Pedersen 2022, Najar 2026), and dual expression of polarization markers is seen in inflamed tissue and thought to contribute to autoimmune disease (Singh 2023). T cell trafficking between blood and gut is distinct from other nonlymphoid organs (Burton 2024), being driven by gut microbiota and inflammatory stimuli (Masopust 2010, Cheroutre 2004, Galván-Peña 2024, Hegazy 2017). Studies of inflamed intestinal tissue in ulcerative colitis (UC) have shown distortion of T cell subsets, increasing regulatory T cells (Tregs) with low immunosuppressive capacity and altered transcriptional profiles of CD8 effector memory (Tem) and Trm cells (Corridoni 2020, Boland 2020).

While T cell migration is an important therapeutic target in IBD and other immune-mediated diseases (Feagan 2013, Sandborn 2005, Sandborn 2021), our understanding of T cell migration across human tissues is limited. Bulk α- or β-chain TCR-seq across human blood and intestinal tissue has demonstrated single-chain sharing across sites with limited resolution (Saravanarajan 2020, Kakuta 2020) and without the power of cell phenotype information (Han 2014). Paired single cell TCR and single cell RNA sequencing (scTCR/RNA-seq) has facilitated the discovery of novel T cell behavior on the basis of function and clonal lineage, including differential Treg plasticity between tonsil and blood (Sureshchandra 2025), de-orphaning of TCR–peptide–HLA ligand pairings in COVID-19 infection (Ford 2023), and distinct clonal origins of granzyme K- and granzyme B-expressing CD8 T cells in rheumatoid arthritis (Dunlap 2024). Luminal contents of the GI tract vary by anatomic site (She 2024, Donaldson 2015, Donaldson 2020, Kulkarni 2025) as do priming tendencies in draining lymphatics (Esterházy 2019), which could plausibly lead to TCR repertoires regionalized by antigen specificity or phenotype. Bulk TCR-seq of the colon has shown high dispersal of CD8 TCR β-chains across colon sites (Williams 2023), although β-chain analysis has been shown to overestimate overlap between distinct repertoires compared to paired αβ TCRs (Dash 2017).

To uncover T cell clonotype localization patterns in health and UC, we apply split-pool barcoding (Parse Biosciences) and flow-cell based barcoding (10x Genomics) to perform scTCR/RNA-seq from human colon and blood. We profiled colon biopsies from eight healthy donors and six donors with active UC. For three healthy donors, we acquired peripheral blood at two timepoints to measure the stability of clonal dispersion across blood and colon. With this combined dataset of 333,088 T cells with paired transcriptome and αβ TCR information (complete T cells), we show that colon and blood are highly compartmentalized, while intra-organ repertoires are largely conserved across time and anatomic distance without distance-based gradients in repertoire overlap. We demonstrate that intra-colon and blood-colon clonal dispersion differs widely among T cell phenotypes, and we identify transcriptional signatures associated with clonal dispersion. Finally, we show that intestinal inflammation in UC alters proportion, transcription, and clonal dispersion of Tregs, CD8 Trms, and CD8 Tems across inflamed sites, perhaps reflecting altered antigenic drivers of T cell expansion and migration. Our findings advance our understanding of T cell surveillance in the gut at homeostasis and in UC.

## Results

T cells with shared antigen specificity can be detected across blood and colon (Hegazy 2017, Martini 2023), and single cell sequencing allows nucleotide-level tracking of clonal populations across tissues coupled with cell phenotypes. We apply this technology to define the extent of TCR sharing between blood and healthy colon, relationship of TCR sharing in the healthy colon to anatomic distance approximated between the sites of tissue sampling, and alterations caused by UC inflammation. We performed ‘deep’ scTCR/RNA-seq with split-pool barcoding on colon and blood from three healthy donors, yielding an average of 19,792 complete T cells per blood sample and 5,435 per colon sample, with high TCR similarity between samples sequenced by both methods (Figure S1A). We performed ‘conventional’ scTCR/RNA-seq with flow-cell barcoding on colon from five healthy donors, yielding an average of 1,284 complete T cells per colon sample (Figure 1A, Table S1). For all healthy donors, we collected biopsies of the cecum (CE), transverse colon (TV), and rectum (RE), which have limited overlap in lymphatic drainage (Denham 2012). T cell trafficking to the colon is modulated by expression of integrin α4β7 (Sandborn 2013). To investigate whether this re-circulating T cell population drives repertoire overlap between blood and colon, we profiled blood at two timepoints and analyzed integrin α4β7 positive and total blood T cell populations separately (Figure S1B). After filtering cells by gene counts, transcript counts, and RNA content, cells were annotated by CellTypist (Domínguez Conde 2022).

**Figure 1.**
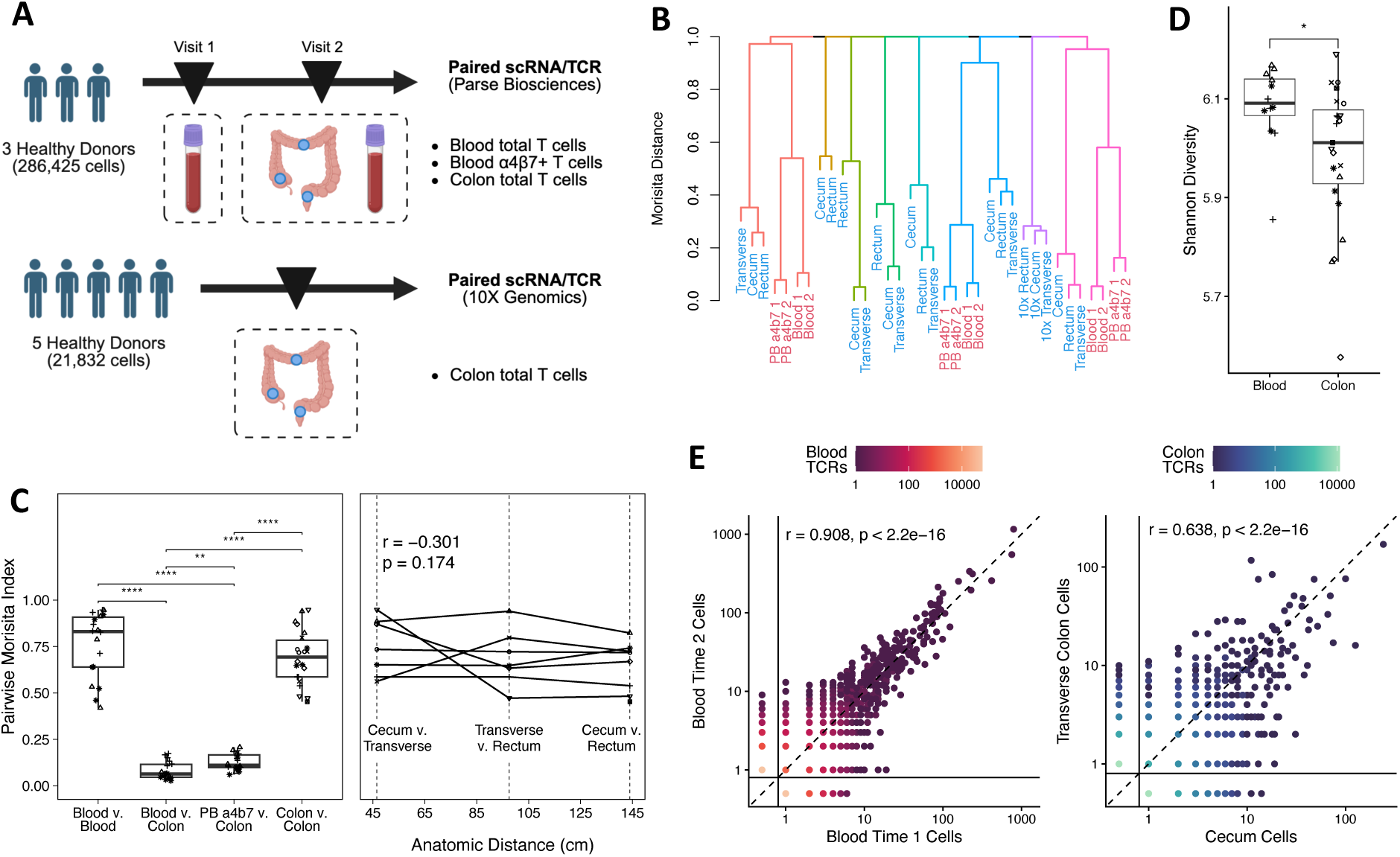
Colon and Blood TCR Repertoires are Compartmentalized and Internally Stable Across Distance and Time. (A) Overview of cohort and experimental design in this study. Healthy donor (n = 8) colons were biopsied at three sites. Three healthy donors were also profiled in autologous peripheral blood sampled at colonoscopy and 1 to 3 weeks before or after. Peripheral blood T cells at all time points were separated into unenriched and integrin α4β7 positive fractions.Created in BioRender. Fischer, J. (2026) https://BioRender.com/j7x81k1 (B) Hierarchical clustering of Morisita distance groups samples by donor and organ. Sample tissue indicated by label color, red indicates blood, blue indicates colon biopsy. Donor (n=8) indicated by branch color. Blood samples enriched for integrin a4b7 or unenriched. (C) Clonal sharing by pairwise Morisita index is high between samples from the same organ and low between colon and blood samples. Enrichment for integrin α4β7 in blood increases clonal overlap with colon. Clonal sharing between colon sites is not significantly associated with anatomic distance. Each point is a comparison between two repertoires, shape represents donor, and lines connect comparisons from same donor. p values for pairwise comparisons calculated by Mann-Whitney U test and corrected for multiple comparisons by Holm method. **p < 0.01, ****p < 0.0001. Correlation and p values calculated by Pearson’s product-moment. (D) Colon tissue is associated with lower TCR diversity than blood by Shannon diversity. All samples (blood n=3, colon n=8) rarefied to lowest sample size. Each point represents one repertoire, shape indicates donor. p value by Mann-Whitney U test. *p < 0.05 (E) Blood clonotypes (n=3 donors) have similar expansion across time points, colon clonotypes (n=8 donors) have similar expansion across cecum and transverse colon. Clonotypes from all healthy donors plotted together, comparisons are restricted to site pairs from the same donor and paired samples are downsampled to equal cell count. Point location indicates number of cells detected at each site per clonotype, point color indicates number of clonotypes with that cell distribution. Clonotypes with 0 cells in one compartment represented as 0.5 (below solid lines) for visualization on log-scale axes, dashed lines indicate equal clone size in both sites. Pearson’s product-moment correlation with p value calculated for each comparison.

### Individual TCR repertoires are unique with modest sharing between blood and colon

To investigate the extent of clonal sharing between blood and colon, we calculated Morisita index, which accounts for amount of shared clonotypes and the similarity of their expansions across two samples, with values ranging from 0 in samples with no shared clonotypes to 1 in highly similar samples (Leick 2020). Hierarchical clustering of samples by Morisita distance grouped each repertoire by subject, demonstrating the uniqueness of each individual’s TCR repertoire. Each donor’s region of the tree was characterized by a tight cluster between blood samples collected over time, tight clustering between samples along the length of the colon, and more distant relationships between the blood and colon samples of each individual (Figure 1B). Within each donor, Morisita index between blood and colon repertoires was significantly lower than between samples of the same organ, with no differences by colon site (Figure 1C, S1C). Integrin α4β7 positive blood showed a small but significant increase in Morisita index with colon compared to unenriched blood (Figure 1C). Blood repertoires did not differ in diversity by integrin α4β7 enrichment (Figure S1D), and colon repertoires showed lower Shannon diversity than blood (Figure 1D), in line with previous reports of oligoclonality in healthy colon (Blumberg 1993, Van Kerckhove 1992). These results suggest that TCR repertoires have near zero overlap between individuals and low overlap between blood and colon with slight enrichment in α4β7 positive blood.

### Intra-colon TCR repertoire sharing is independent of inter-site distance

To investigate the extent of clonal compartmentalization across the length of the colon, we compared TCR repertoires across three colon sites from 8 healthy donors. The TCR repertoire overlap across colon sites was heterogeneous among donors. Repertoire overlap was not associated with anatomic distance between sampling sites, as the comparison with the shortest distance (cecum vs. transverse colon) showed similar overlap to the comparison with the longest distance (cecum vs. rectum) (Figure 1C). Repertoire overlap was also not associated with donor (Figure S1E), age, or sampling depth (Figure S1F). For blood-to-blood comparisons, clonotype abundance was strongly correlated across timepoints (Pearson r = 0.908; p<2.2x10^16^; Figures 1E, S1G, Table S2). Clonotype abundance was also correlated between colon sites (CE-TV, Pearson r = 0.638, p<2.2x10^16^; CE-RE r = 0.523, p<2.2x10^16^; TV-RE r = 0.536, p<2.2x10^16^) (Figure 1E, S1H, Table S2). Our data indicate that clonotypes are highly distributed among colon sites, suggesting shared antigen reactivity across anatomic distance rather than curation toward locally abundant antigens.

### CD8 Tem show high blood-colon dispersal, while Tregs show clonal restriction

To determine whether the 2-11% of clonotypes that we found shared between blood and colon was influenced by cell phenotype (Figure S2A), we measured the dispersion of T cells with identical αβ V(D)J recombination across compartments for each phenotype. We individually measured repertoire overlap between all colon sites and blood in three healthy donors. Blood and colon T cells showed large differences in phenotype, with a near absence of effector CD4 cell phenotypes in blood (Figure 2A). We measured the evenness of clonotype dispersion between blood and colon using Shannon entropy per clonotype weighted by clonal size as previously described (Zhang 2018), which we refer to as dispersion index. Pairwise dispersion index was calculated for each phenotype subset across each pair of intra-donor blood and colon samples. Pairwise dispersion index ranges from 0 (total clonotype segregation) to 1 (perfect clonal mixing). Among cell phenotypes with over 100 cells detected in each compartment cumulatively across all donors, CD8 Tem clonotypes were most highly dispersed between blood and colon, suggesting frequent interchange or independent expansion toward shared antigens, while Tregs showed uniquely low dispersion, suggesting distinct antigen reactivity between blood and colon (Figure 2B). CD4 Tcm cells showed higher clonal sharing than Treg, suggesting higher exchange of conventional T helper cells over Tregs between blood and colon (Figure 2B). Inter-organ clonotype abundance had a low but significant correlation across organs (Pearson r = 0.096, p<2.2x10^-16^; Figure 2C) relative to intra-blood and intra-colon comparisons (Figure 1E), with most clonotypes showing a bias toward expansion in one compartment. Our findings demonstrate a consistent pattern of blood-colon sharing of CD8 Tem clones and support a model of Treg clonal compartmentalization between blood and colon, possibly mediated by selection for microbial reactivity.

**Figure 2.**
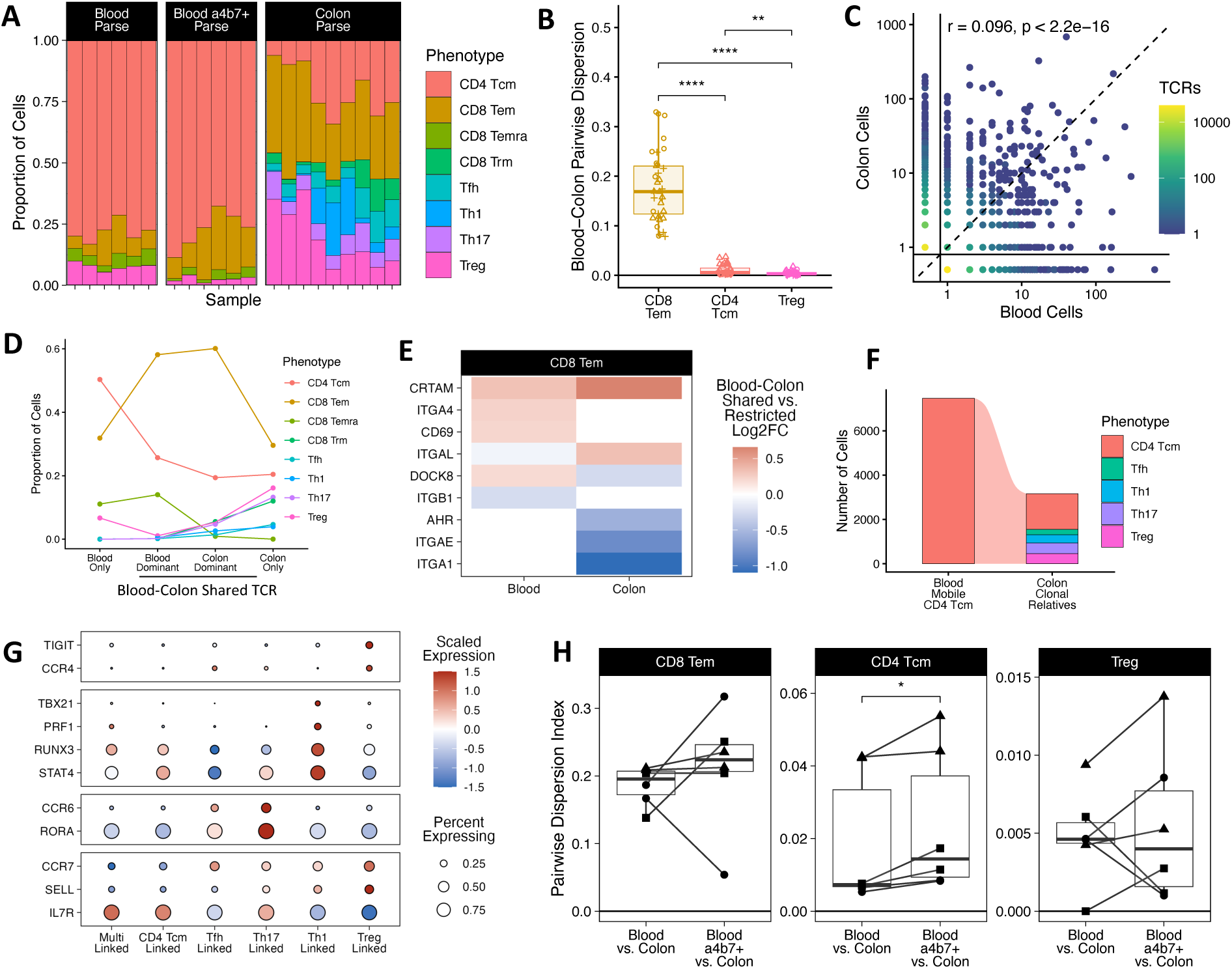
Clonotype Mobility Between Colon and Blood Varies by Phenotype. (A) T cell population phenotypes vary between blood and colon. (B) Cytotoxic T effector memory compartment shows higher clonal migration between blood and colon than all other phenotypes. Pairwise dispersion indicates evenness of T cell clones across two sites with matching phenotype labels. Each point represents a comparison between one blood and one colon repertoire, shape indicates donor (n=3). p values calculated by Mann-Whitney U test. **p < 0.01, ****p < 0.0001 (C) Blood and colon clonotype abundance (n=3 donors) is weakly correlated across organs. Clonotypes from all donors plotted together, comparisons are restricted to site pairs from the same donor. Point location indicates number of cells detected at each site per clonotype, point color indicates number of clonotypes matching given cell distribution. Clonotypes with 0 cells in one compartment represented as 0.5 (below solid lines) for visualization on log-scale axes, dashed lines indicate equal clone size in both sites. Correlation and p value calculated by Pearson’s product-moment. (D) Phenotype of T cells within clonotype dispersion categories. Clonotypes with greater number of cells in blood or colon termed “blood dominant” or “colon dominant”, respectively. (E) Differentially expressed genes of CD8 Tem cells in inter-organ mobile clonotypes relative to cells in single organ clonotypes. Included genes meet significance cutoff of FDR = 0.05. (F) Phenotype identies of CD4 T cells in colon with clonal relationships to circulating CD4 Tcm cells across healthy donors (n = 3). (G) Differential gene expression of polarization markers within blood CD4 Tcm by phenotype of colon-linked cells. Cells belonging to clonotypes with multiple colon CD4 phenotypes labeled “Multi Linked”. Selected genes meet significance cutoff of FDR = 0.05. Color indicates log1p-transformed normalized transcript count Z-scaled scaled by gene, size indicates proportion of cells in group with nonzero expression of gene. (H) Pairwise migration between blood and colon CD4 Tcm cells is higher among integrin a4b7 positive blood than unenriched blood. Each point represents comparison between pooled colon sites and blood at one time point, shape indicates donor. p value by Wilcoxon signed rank test. *p < 0.05

### Cross-compartment clonal relationships reveal transcriptional signatures of migration and polarization state

In addition to measuring clonal dispersion by phenotype, scTCR/RNA-seq allows us to discover possible transcriptional drivers of dispersion. To characterize cell-level function within mixed or restricted clonal populations, we measured differential gene expression (DGE) by Wilcoxon rank-sum test with p value correction by Benjamini-Hochberg procedure for cells belonging to clonotypes that were shared across blood and colon versus cells belonging to clonotypes restricted to one compartment. After filtering cells belonging to expanded clonotypes (>= 2 cells across >=1 sites), we subset cells by compartment to exclude transcriptional differences due to tissue processing. We further subset cells by phenotype to reduce the influence of imbalanced phenotype representation in dispersed clonotypes (Figure 2D). Within the CD8 Tem subset, we identified 507 differentially expressed genes between shared and exclusive clones in blood and 430 differentially expressed genes between shared and exclusive clones in colon (Table S3). Among these genes, shared CD8 Tem clones in colon showed downregulation of tissue residency-associated markers (*ITGA1, ITGAE, AHR*), while shared CD8 Tem clones in blood showed upregulation of tissue homing and residency genes (*ITGA4, CD69, DOCK8*) relative to the single site restricted Tem cells (Figure 2E). The expression of the activation marker *CRTAM* was upregulated in cross-compartment clonal groups in both tissues, suggesting a role in trafficking into and out of tissues consistent with a previous report of lower tissue infiltration and lymph node retention in *Crtam*^-/-^ mice (Takeuchi 2009). These findings suggest that colon CD8 Tem populations related to blood are less adapted to tissue residency, possibly due to recent arrival or ongoing emigration at the clonal population level.

Microbe-reactive peripheral blood CD4 memory T cells show a diversity of effector states when re-stimulated (Martini 2023, Hegazy 2017). Given that our dataset showed quiescent CD4 Tcm as the dominant circulating conventional CD4 T cell population, we investigated how blood Tcm were related to effector populations in the colon (Figure 2D). We subset blood CD4 Tcm with clonal relative in colon and performed DGE analysis based on the effector phenotypes of colon relatives (Figure 2F). Circulating CD4 Tcm linked to Treg clones in the colon upregulated *TIGIT* and *CCR4*, those linked to Th1 clones upregulated *TBX21, PRF1, RUNX3,* and *STAT4*, while those linked to Th17 upregulated *CCR6* and *RORA*. (Figure 2G, Table S4). In addition, *IL7R* was upregulated by Th17-linked cells and downregulated by Treg-linked cells, in line with prior reports of Th17 but not Treg dependence on IL-7 signaling (Meyer 2022). Taken together this suggests that clones of the same lineage conserve polarization state between blood and colon. The dispersion index for CD4 Tcm was uniquely elevated in integrin α4β7 positive blood compared to unenriched blood, suggesting a greater homing effect of integrin α4β7 in CD4 Tcm (Figure 2H). Colon repertoires shared more clonotypes with blood than blood did with colon, and cells belonging to shared clonotypes made up a larger share of colon than blood, suggesting migration occurs primarily from blood to colon (Figure S2A, S2B). We found that circulating CD4 Tcm cells transcriptionally resemble colon CD4 effectors with shared TCRs and that integrin α4β7 enriches clonal sharing between blood and colon within the CD4 Tcm compartment but not CD8 Tem or Treg.

### Th17 are more dispersed along the colon than Treg, and highly dispersed clones upregulate residency markers

Given the high rate of clonal sharing across the colon, independent of anatomic distance (Figures 1C, 1E), we sought to determine whether cell phenotype influences clonal sharing. We measured dispersion index of phenotype groups across distant colon sites from 8 healthy donors (Figure 1A). Colon samples showed similar proportions of major phenotypes across library construction methods (Figure S3A). Similar to blood-gut sharing, clonotype migration across colon sites was significantly higher in cytotoxic T cells (Mann-Whitney U test, p = 1.9 x 10^-12^; Figure 3A), and cytotoxic T cells made up a significantly greater fraction of clonotypes detected at all three colon sites (Wilcoxon signed-rank, p = 0.032; Figure 3B). Among CD4 phenotypes detected in both library preparation methods, we found that Th17 cells showed the greatest pairwise dispersion, while Tregs showed the least (Figure 3C). Within each phenotype, DGE analysis showed cells belonging to expanded clonotypes detected at multiple sites had the highest expression of residency-associated genes (*ITGAE, ITGA1, AHR*), while cells belonging to singleton clonotypes had the highest expression of migration genes (*CCR7, ITGB2, CXCR4*) (Figure 3D, Table S5), suggesting the clonotypes with the largest footprint in the colon are less migratory. Low migration of widely shared clones may suggest the lamina propria has a limited window of tissue seeding during systemic immune activation, as in the intraepithelial compartment (Masopust 2010). Taken together, this demonstrates that Th17 clones are more widely dispersed than Treg clones, and that wide intra-colon dispersion is associated with residency signatures rather than mobility.

**Figure 3.**
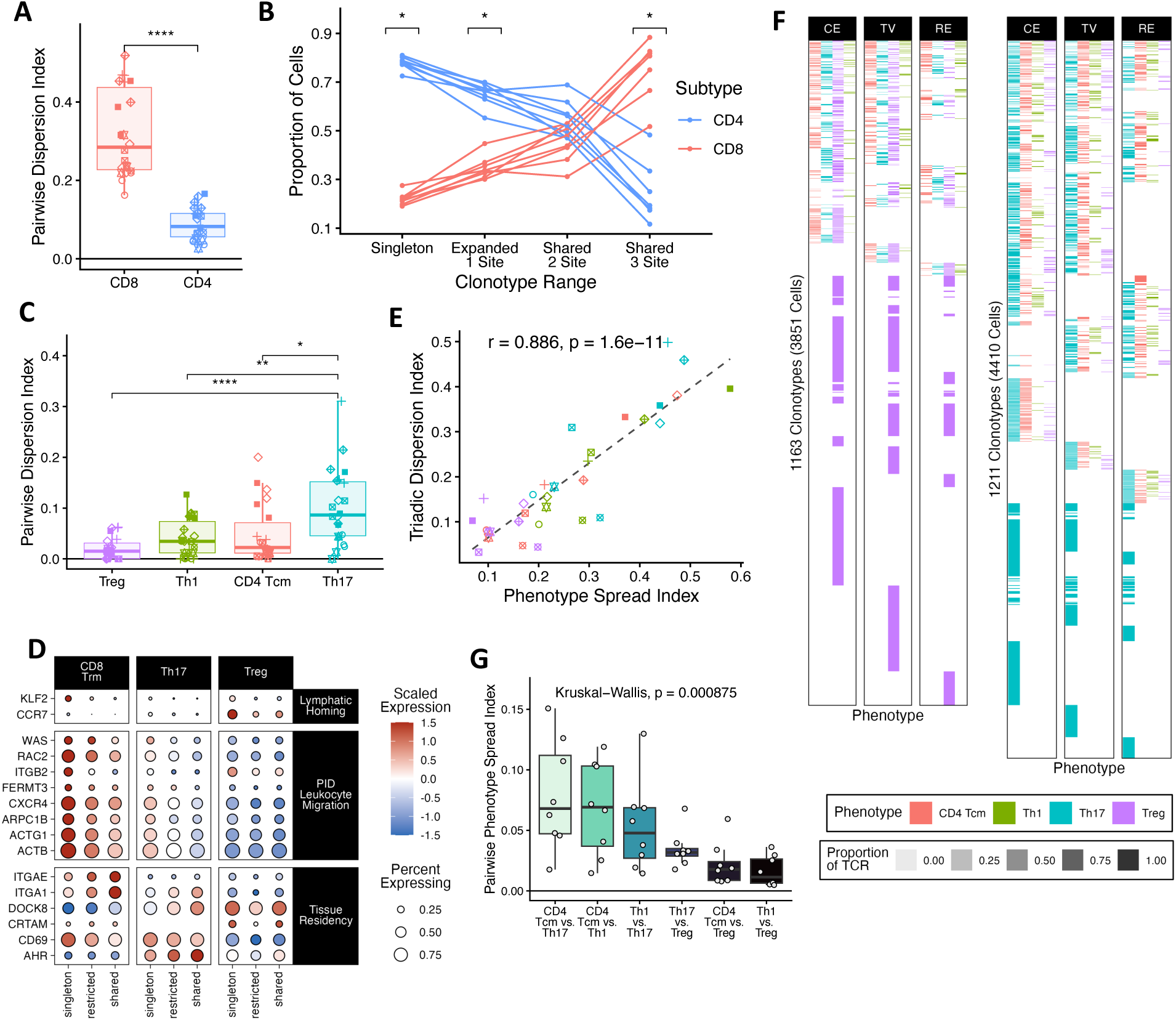
Clonotype Mobility Between Colon Sites Varies by Phenotype. (A) Pairwise dispersion index is higher in CD8 T cells than CD4 T cells. Dispersion index indicates evenness of clonal T cell spread across sites with matching phenotype labels. p values calculated by Mann-Whitney U test. ****p < 0.0001 (B) CD4 T cells make up a greater proportion of clonotypes detected at one site, and CD8 T cells make up a greater proportion of clonotypes detected at three sites. Individual points represent total from each donor (n = 8), with lines connecting measurements from same donor. p values calculated by Wilcoxon signed-rank test and corrected for multiple comparisons by Holm method. *p < 0.05 (C) Tregs have lower pairwise mobility across colon compared to Th17. p values calculated by Mann-Whitney U test and corrected for multiple comparisons by Holm method. *p < 0.05, **p < 0.01, ****p < 0.0001 (D) Differential gene expression of mobility and residency markers by clonal expansion and site-site sharing within each phenotype. Cells belonging to clonotype with one cell termed “singleton”, cells belonging to clonally expanded clonotypes detected at one site termed “restricted”, cells belonging to clonotypes detected at multiple sites termed “shared”. Included genes meet significance cutoff of FDR < 0.05. (E) Clonotype dispersion is strongly associated with phenotypic diversity. Phenotype spread index measures evenness of clonally related cells across phenotypes. Correlation and p value calculated by Pearson’s product-moment. (F) Heatmap of expanded Treg and Th17 clonotypes and all CD4 T cells with shared TCR across colon sites. Each row represents a clonotype, vertical facets separate each colon site, column and color represents each phenotype label, transparency indicates proportion of T cells within a clonotype. Clonotypes from all donors are plotted together. (G) Pairwise phenotype spread index between CD4 phenotypes in colon. Each point represents comparison between two phenotypes for pooled sites from same donor. p value calculated by Kruskal-Wallis.

### T cell clonal groups with high intra-colon dispersion are phenotypically varied

Prior studies have demonstrated that targeted microbial and environmental antigens induce phenotypically diverse T cell responses in humans (Zielinski 2012, Martini 2023, Hegazy 2017), including reciprocal Treg and conventional T helper responses (Mucida 2007, Gagliani 2015, Bacher 2016, Bacher 2014, Bacher 2013, Xu 2018). To address the question of clonal phenotype diversity in the healthy colon, we measured the phenotype spread index of clonotypes across phenotypes as previously described (Zhang 2018). Similar to the dispersion index, the phenotype spread index is calculated as a weighted average of Shannon entropy of cells distributed across phenotypes for each TCR in a sample. A phenotype spread index value of 0 indicates no phenotype diversity among clonally related T cells, while higher values suggest more balanced phenotype variation. For each CD4 T cell phenotype, the phenotype spread index was correlated with dispersion index (Pearson r = 0.886, p=1.64x10^-11^; Figure 3E, Table S2), suggesting clonotypes that are broadly distributed are most likely to contain a greater breadth of phenotypes (Figures 3F, S3C). Phenotype spread index between phenotype pairs varied significantly (Kruskal-Wallis test, p = 0.001, Figure 3G), with the highest pairwise phenotype spread index values between Th17, Th1, and CD4 Tcm, similar to phenotype variation in clonal groups across blood and colon (Figures 2F, 3G). Taken together, this suggests that as T cell clonal groups become more ubiquitous in the colon, their members acquire a greater breadth of effector functions, possibly reflective of localized signaling differences in the lamina propria microenvironment.

### Inflamed UC colon sites have elevated Tregs and increased clonal dispersion of CD8 Tem

To assess how UC inflammation alters T cell phenotypes and clonal sharing across multiple sites of the colon, we profiled T cells from inflamed and uninflamed colon tissue from six active UC patients, including three donors with rectal or left colon inflammation and a ‘cecal patch’ (Fard 2022), separated by uninflamed transverse colon (Figure 4A, Table S1). While UC typically begins in the rectum with proximal progression, these three patients had endoscopically normal colon tissue between inflamed sites, suggesting independent inflammatory triggers at these sites. To minimize technical differences in phenotype annotation and transcription, we compared UC donors to healthy donors that were barcoded by the same 10X method. Inflamed colon tissue showed significantly elevated proportions of Tregs and decreased proportions of CD8 Trms (Figure 4B, S4A), in line with previous scRNA-seq findings (Boland 2020, Smillie 2019). To examine transcription differences between inflamed and noninflamed tissue, we measured differential gene expression of complete T cells of the same phenotype between UC inflamed, UC uninflamed, and healthy donor samples by Wilcoxon rank-sum test with p value correction by Benjamini-Hochberg procedure. T cells in inflamed tissue showed upregulation of exhaustion markers (*LAG3, ENTPD1, CTLA4*), upregulation of inhibitory proteins (*NKFBIA, PRDM1*), and downregulation of AP-1 components (*JUN, FOS*), suggesting chronic simulation of all T cell subsets (Figure 4C, Table S6). Phenotype-specific differences include upregulation of *IL2RA* in Tregs, *GNLY* and *GZMB* in CD8 Tem and Trm, and *IL17A* and *IL22* in Th17 (Figure 4C, Table S6). Downregulation of *IL2* in CD4 Tem, Th17, and Th1 suggests downregulation of inflammatory signaling due to exhaustion or suppression by Tregs (Figure 4C, Table S6). Comparing our inflamed Treg differentially expressed genes to those published by Boland et al, we identify a common signature of UC inflamed Tregs (Figure S4B) including upregulation of immunosuppressive genes (*ENTPD1*, *LAG3*, *TNFRSF4*).

**Figure 4.**
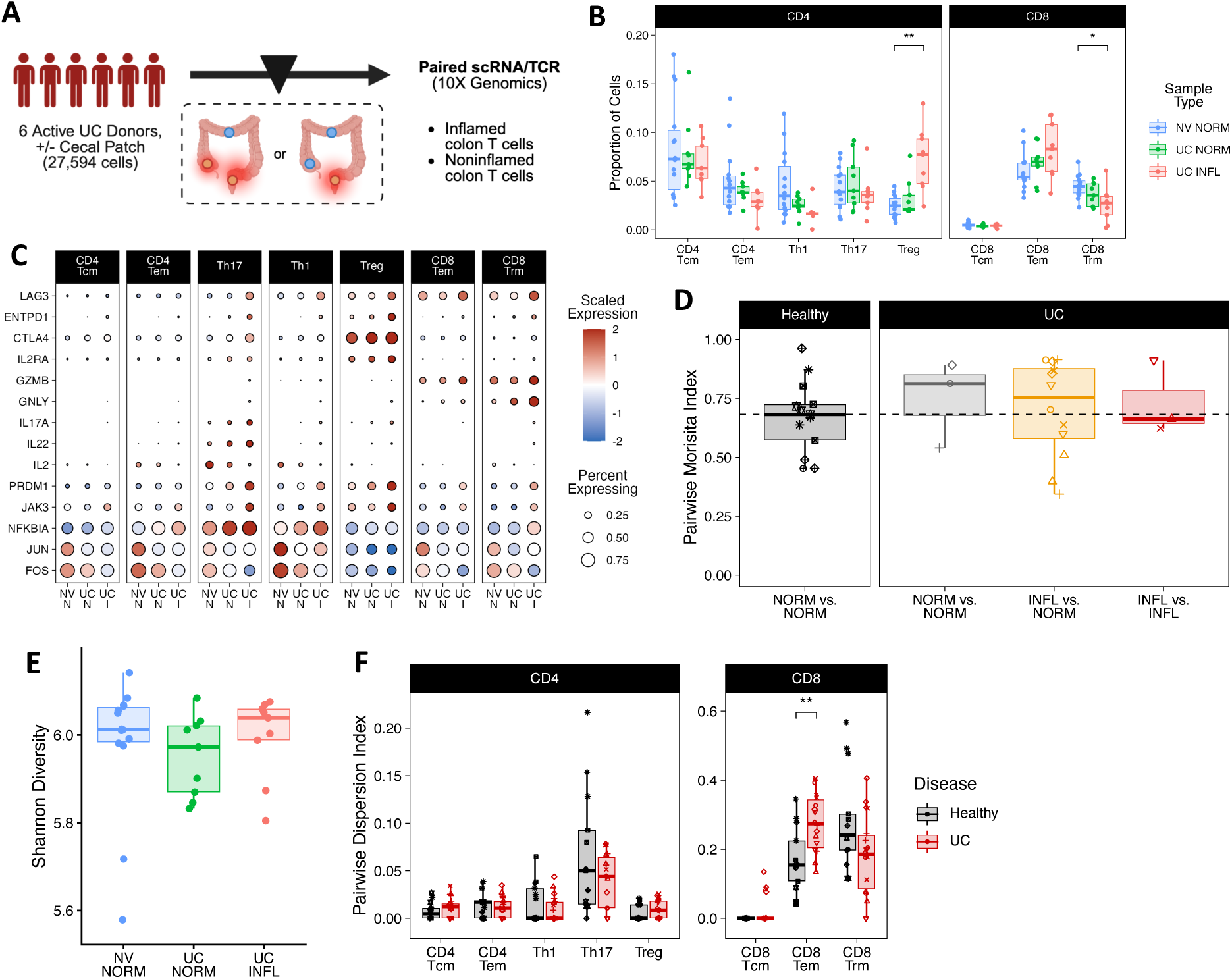
Clonally Related Tregs are Elevated and Shared Across Inflamed Colon Sites. (A) Overview of biopsy collection from donors with active UC (n = 6). Three patients had discontinuous inflammation of distant anatomic sites separated by macroscopically normal tissue (two inflamed sites and one noninflamed site). Three patients had one inflamed site and two noninflamed sites. Created in BioRender. Fischer, J. (2026) https://BioRender.com/j1vj6op (B) Proportion of Tregs out of helper T cells is elevated in inflamed colon. CD8 Trms are reduced in inflamed colon. p values calculated by Mann-Whitney U test and corrected for multiple comparisons by Holm method. *p < 0.05, **p < 0.01 (C) Bubble plot of differentially expressed genes within T cell phenotypes between inflamed (UC I), UC noninflamed (UC N), and healthy noninflamed (NV N) tissue. Included genes meet significance cutoff of FDR = 0.05. (D) Colon site-site morisita index does not vary between healthy and UC. p values calculated by Mann-Whitney U test and corrected for multiple comparisons by Holm method. **p < 0.01, ****p < 0.0001 (E) Colon TCR diversity does not vary between healthy, UC inflamed, and UC noninflamed tissue by Shannon entropy. All samples (healthy n=5, UC n=6) rarefied to lowest sample size. p value by Mann-Whitney U test. (F) Pairwise dispersion in CD8 Tems is higher in UC donors. Each point represents comparison between two sites from same donor after rarefying to equal sample size, shape represents donor. p values calculated by Mann-Whitney U test and corrected for multiple comparisons by Holm method. **p < 0.01

By comparing shared clonotypes between inflamed-inflamed, inflamed-noninflamed, or noninflamed-noninflamed, we can gain insights into if the immune changes above represent a broad non-specific response or potentially expansions and depletions of specific clonotypes. The combined T cell site-to-site repertoire overlap measured by Morisita index was not different between UC and healthy (Figure 4D). Similarly, overall TCR repertoire diversity measured by Shannon index was not significantly different between healthy, UC noninflamed, and UC inflamed (Figure 4E, S4C), which is consistent with single cell TCR-seq colon results (Boland 2020).

To investigate whether clonal sharing was altered by inflammation in a phenotype-specific manner, we calculated pairwise dispersion index by phenotype after rarefying to the minimum population size per donor to reduce the impact of differential abundance of phenotypes with inflammation. When grouped by disease, pairwise dispersion index was significantly elevated in CD8 Tems in UC donors compared to healthy, suggesting UC promotes the clonal homogeneity in this compartment (Figure 4F). When site-site comparisons were split by inflammation status, Tregs showed a trend toward increased clonal sharing among inflamed sites, possibly indicating shared antigenic targets across distant inflamed sites (Figures S4D). In contrast, CD8 Trm populations trended toward lower dispersion across inflamed sites, suggesting UC inflammation interferes with intra-colon clonal ubiquity in this compartment (Figure S4D). Surprisingly, inflamed tissue sites showed trends toward reductions in phenotype spread index among CD4 Tcm, CD4 Tem, and Th17 phenotypes (Figure S4F).

Taken together, this suggests UC inflammation induces T cell exhaustion signatures while leaving the TCR repertoire mostly unchanged from uninflamed colon. We observed elevated CD8 Tem clonal dispersion in UC donors, but differences did not correspond to tissue inflammation. While we did not detect large clonal expansion of pathogenic clonotypes specific to inflamed sites in UC, our findings suggest that Tregs at distant inflamed sites have a higher degree of clonal sharing. Treg clonal sharing could be due to local expansion of pre-existing shared clonotypes or recruitment of the same clones from the periphery in response to shared antigens in the inflamed environment.

## Discussion

In this study, we utilize two methods of single cell TCR profiling to study T cell clonal dynamics in the colon in health and ulcerative colitis. Our use of split-pool barcoding allowed us to profile a greater number of paired TCRs across blood and colon than previous scTCR/RNA-seq studies in humans (Boland 2020, Zhang 2018, Thomas 2024) and reveal dynamics of CD4 T cell migration between blood and colon that were previously unexplored by bulk TCR-seq (Williams 2023).

Our results demonstrate that clonal sharing between blood and colon is consistently lower than either tissue with itself across time or distance. Further, we find a consistent pattern of blood-colon sharing of CD8 Tem clones contrasted by colon Treg clonal compartmentalization from blood as has been observed in mice (Burton 2024, Lathrop 2011), possibly due to differences in induced versus thymic origin and mediated by selection for microbial reactivity (Gu 2024). Bacterial and fungal reactive CD4 T cells have been detected in peripheral blood and colon in healthy and IBD donors, mainly derived from central memory populations (Martini 2023, Hegazy 2017, Pedersen 2022), suggesting ongoing exchange of microbe-reactive colon T cells with blood. Our results demonstrate that in addition to shared antigen reactivity, a subset of circulating T cells bears identical TCRs to colon effector populations and upregulate corresponding polarization markers.

We find that the TCR repertoire of the gut is highly shared independent of anatomic distance (similar to blood over time), particularly among cytotoxic and Th17 clones. Tregs show greater regional differences in TCR repertoire. In addition, we show that the extent of a clonotype’s distribution in the colon is positively associated with phenotypic breadth of its constituent T cells. This suggests that as descendants of a common T cell progenitor spread and encounter new tissue environments, they adapt their effector function. TCR-agnostic flow cytometry studies have shown elevated rates of T cells with dual expression of IL-17 and FOXP3 or IFN-γ in blood and colon of IBD patients, (Ueno 2018), suggesting plasticity of Th17 cells may contribute to disease. We observed a trend of lower phenotype crossover of CD4 clonal populations within inflamed colon compared to healthy, consistent with a recent scTCR study demonstrating microbiota-induced T cell clones take on a more limited range of phenotypes in tumor compared to intestine (Najar 2026), and consistent with the notion that polarization state is the result of environmental cues following TCR signaling (Zhou 2009, Pedersen 2022). We find that widely distributed intra-colon T cell clones are associated with upregulated residency markers, suggesting dispersion across the colon is not mediated by active cell migration and may reflect prior migratory events. These findings align with previous studies demonstrating limited windows of barrier tissue T cell seeding from blood during infection independent of the infected tissue (Masopust 2010, Cheroutre 2004). In contrast, we demonstrate that clonotype sharing between blood and gut is associated with increased expression of migration markers, suggesting the lower level of TCR sharing between blood and colon is due to active migration of minor clonal populations.

We observed an elevation of Tregs and depletion of CD8 Trms in active UC lesions, similar to prior studies (Boland 2020, Holmén 2006). We observed upregulation of T cell exhaustion markers (*CTLA4, ENTPD1, LAG3*) in inflamed colon, consistent with other single cell studies of UC (Uzzan 2022, Boland 2020, Corridoni 2020). While overall intra-colon repertoire overlap was similar between UC and healthy colon, CD8 Tem populations showed greater dispersion across sites in UC, suggesting altered clonal dynamics in this subset.

Consistent with prior TCR profiling of inflamed colon tissue from UC patients (Boland 2020, Werner 2018), overall repertoire diversity was not reduced by inflammation. Microbe-specific CD4 T cell transfer has been shown to mediate autoinflammation in disease models (Xu 2018, Zeng 2023), and endogenously generated microbe-reactive CD4 T cell populations are diverse at the β-chain CDR3 or V/J gene level (Hegazy 2017, Martini 2023, Xu 2018, Bacher 2019), suggesting antigen-specific CD4 T cell response in UC may be the sum of contributions from a large number of clonal populations.

Taken together, our study highlights structural principles of the human colon TCR repertoire in health and UC. We identify that colon repertoires are highly dispersed across large anatomic distance with the exception of Tregs, which are compartmentalized both from other colon sites and from peripheral blood. By coupling transcription to αβ TCRs, we were able to identify opposing mobility signatures in highly dispersed blood-colon and intra-colon clones, supporting different mechanisms of clonal dispersion. Our findings in UC highlight the importance of resolving the TCR repertoire by T cell phenotype, which reveals differences in clonal dynamics not detectable in aggregate repertoires. Further study is needed to assess the response of inflamed clonal populations in IBD to treatment, their contribution to disease remission, and their antigenic targets.

## Methods

### Study design

This study was designed to profile the localization of T cells along sites of the colon in health and UC. We generated and analyzed a scRNA/TCR-seq dataset from human colon lamina propria and peripheral blood.

### Human participants

Institutional research board (IRB, STUDY-17-01304-MOD0043) approval for blood and biopsy collection was obtained at Mount Sinai Hospital. Colon tissue and peripheral blood were obtained from patients undergoing colonoscopy at Mount Sinai Hospital, IBD Center, after obtaining informed consent. Healthy donors underwent colonoscopy for routine colorectal cancer screening or noninflammatory gastrointestinal symptoms. Inclusion criteria included minimum patient age of 18 years, absence of significant comorbidities, and active disease in UC donors. Characteristics of included participants are available in Table S1.

### Human peripheral blood mononuclear cell isolation

Three healthy controls had 20 mL peripheral blood drawn during colonoscopy and at one additional time point at least one week before or after biopsy collection. Blood was collected in EDTA-coated tubes and diluted with 2 volumes HBSS. Diluted blood was overlaid onto ½ volume Percoll (Cytiva) before centrifugation for 30 minutes at 500g. Mononuclear cells were collected from the interface, washed with 4-5 volumes of HBSS, and centrifuged for 10 minutes at 500g. Red blood cells were lysed by resuspension in ACK lysis buffer and incubation at room temperature for 5 minutes. Cells were washed with HBSS once more and resuspended in 1 mL MACS buffer (PBS with 10 mM HEPES, 5 mM EDTA, and 0.1% FBS).

### Human colon cell isolation

From donors processed by Parse Biosciences library preparation, eight to twelve pinch biopsy samples were taken from cecum, transverse colon, and rectum during colonoscopy. Biopsies from the same site were placed in 50 mL tubes of RPMI on ice and immediately transported for dissociation and digestion. Tissue fragments were transferred to 50 mL tubes containing 20 mL of predigestion buffer (HBSS without Mg/Ca, 10 mM HEPES, 5 mM EDTA, 1 mM DTT, 5% FBS), shaken horizontally at 180 rpm at 37°C for 20 minutes, and vortexed at 3000 rpm for 1 minute. Shaking incubation and vortexing were repeated once in predigestion buffer and once in wash buffer (HBSS with 10 mM HEPES). All tubes were kept on ice between incubations.

Epithelium-dissociated tissue was then digested at 37°C with LP Dissociation Kit (Miltenyi), gentleMACS C Tubes and gentleMACS Octo Dissociator (Miltenyi) set to ‘37C_m_LPDK_1’ program. Cell suspensions were triturated by 18G needle and syringe, passed through 100 μm filter, and washed in MACS buffer. Cells were centrifuged at 450g for 10 minutes at 4°C and resuspended in 350 μL MACS buffer. Lamina propria cell suspensions were enriched for live cells using Dead Cell Removal Kit (Miltenyi 130-090-101) over LS Columns (Miltenyi 130-042-401) on a quadroMACS magnetic rack (Miltenyi).

For donors processed with 10x Genomics only, biopsies were transferred to dissociation buffer (HBSS without Mg/Ca, 10 mM HEPES, 5 mM EDTA, 10% heat-inactivated FBS), shaken vertically at 37°C at 110 rpm for 20 minutes, and vortexed at 70% max speed for 10 seconds to release epithelium. Incubation and vortexing in dissociation buffer was repeated for a total of two washes, followed by enzymatic digestion (RPMI, 2% FBS, 0.5 mg/mL Collagenase IV (Sigma), 0.1 mg/mL DNase I (Roche)) with vertical shaking at 37°C at 180 rpm for 40 minutes. Post-digestion tissue was vortexed for 30 seconds at max power and triturated with 18G needle and syringe. Cell suspensions were passed through 100 µm and 40 µm filters in sequence followed by dead cell removal using Dead Cell Removal Kit (Miltenyi 130-090-101) following manufacturer’s instructions.

### Magnetic T cell enrichment

PBMCs and live cell enriched lamina propria suspensions were positively selected for T cells using Human CD3 MicroBeads (Miltenyi 130-050-101) and LS Columns (Miltenyi 130-042-401) over quadroMACS magnetic rack.

### Flow cytometry and cell sorting

A subset of CD3 enriched PBMCs from each timepoint was incubated with conjugated antibodies in FACS buffer (PBS with 10 mM HEPES, 5 mM EDTA, and 0.1% FBS) for 30 minutes at 4°C covered from light. Dead cells were excluded with DAPI staining. The following antibodies were diluted 1:100 and used for surface staining: Integrin α4β7-PE (Biotechne FAB10078P), CD19-PE-Cy7 (Biolegend 302216), CD38-APC (BD Biosciences 560980), CD3-AF488 (Biolegend 317310), CD45-BV650 (Biolegend 304044), CD8-Percp/5.5 (Biolegend 344710), CD4-BV421 (Biolegend 344632). Cells positive for integrin α4β7 were sorted using a BD FACSAria II instrument using standard protocols and gating strategy shown in Figure S1B.

### Parse Biosciences library preparation and sequencing

T cell enriched cell suspensions were immediately fixed and permeabilized using Evercode v3 Cell Fixation kits (Parse ECFC3300) and frozen at −80°C with controlled cooling using a Mr. Frosty Freezing Container (Thermo Scientific). Samples were stored at −80°C for no more than 6 months prior to barcoding with Evercode v3 Human WT + TCR Mega kit (Parse ECIT2500).

Samples were thawed, counted, diluted, and barcoded in a single batch with a target output of 1 million barcoded cells captured in scRNA-seq library. Libraries were sequenced on an Illumina NovaSeq instrument. TCR libraries were generated according to manufacturer’s instructions and sequenced on an Illumina NovaSeq instrument.

### 10x Genomics library preparation and sequencing

Unselected cell suspensions were processed into single cell RNA-seq and TCR-seq libraries according to manufacturer instructions for 10x Genomics Chromium Single Cell 5’ Gene Expression and V(D)J Amplification kits. Colon sites from same donor were labeled by hash tagging and pooled prior to barcoding. Gene expression and TCR libraries were sequenced on Illumina instruments.

### Single-cell RNA-seq

Reads from scRNA-seq libraries were aligned to reference genome hg38 and processed by unique molecular identifier (UMI) into transcript counts using Split-Pipe version 1.4.1 (Parse Biosciences) or Cell Ranger version 7.0.1 (10x Genomics) with default parameters. Gene expression data was loaded into Python version 3.13.7 with scanpy version 1.11.4 and filtered for cells with under 5,000 genes, under 25,000 transcripts, and under 25% mitochondrial transcripts. UMI-based transcript counts were normalized with ‘pp.normalize_total’ to target sum of 10,000 and scaled with ‘pp.log1p’. Differential gene expression (DGE) was assessed by Mann-Whitney U test with p value correction by Benjamini-Hochberg using the sc.tl.rank_genes_groups function of scanpy. Genes with an adjusted p-value < 0.05 were considered differentially expressed.

### CellTypist Phenotype Annotation

CellTypist version 1.7.1 was used to assign phenotype labels to cells in scRNA-seq dataset. Filtered, normalized, and log1p-transformed transcript count data was used as input to the ‘celltypist.annotate’ command with majority voting using the ‘Immune_All_Low.pkl’ version 2 reference library of low-hierarchy immune cell types (Domínguez Conde 2022). Cells with consensus T cell annotation were paired with scTCR data for repertoire subset analysis.

### TCR Repertoire Analysis

Reads from TCR-seq libraries were processed with Split-Pipe version 1.4.1 or Cell Ranger version 7.0.1, producing a filtered AIRR format file. Annotated T cells were linked to TCR data by cell barcode to produce 333,088 complete T cells. T cell transcriptomes were linked to paired αβ TCRs at a rate of 60.6% for Parse libraries and 64.2% for 10x libraries. One donor was profiled by both split-seq and 10X with similar repertoires across methods (Figure S1A). For all TCR repertoire analyses, clonotypes are defined as productive paired αβ CDR3 nucleotide sequences. Cells with only alpha or beta chain CDR3 information were excluded from analysis. Cells with more than one alpha or beta chain were assigned to the chain with the higher transcript count. Clonal overlap was assessed using Morisita distance using the R package vegan v2.7-2 and the ‘vegdist’ command on the count matrix of clonotypes by samples. The resulting distance matrix was used to generate the global repertoire similarity dendrogram using the base R ‘hclust’ command with “complete” agglomeration method and colored with R package dendextend v1.19.1. Pairwise Morisita distance was extracted from the distance matrix for each pair of samples from the same donor. Morisita index was calculated as 1-Morisita distance. Shannon diversity was calculated on the count matrix for each sample using the vegan command ‘diversity’. Anatomic distances between gut sites were estimated from published averages of the human colon (Denham 2012).

The STARTRAC (Zhang 2018) algorithm was adapted to analyze clonal relationships across sites. Briefly, the dispersion index, based on STARTRAC-migr metric and renamed to emphasize population distribution over cell mobility, is a measure of how evenly cells with matching TCRs and phenotypes are distributed across body sites using Shannon entropy. This index is calculated for each clonotype, weighted by the number of cells per clonotype, and averaged for each phenotype. The pairwise dispersion index is calculated across each unique combination of sample pairs, and the triadic dispersion index is calculated across three samples. The phenotype spread index, based on the STARTRAC-tran metric, measures weighted Shannon entropy across phenotypes for cells with shared TCRs. Pairwise phenotype spread index was calculated on pairs of phenotypes across all sites within a donor.

For classification of inter-organ clonotype sharing, all clonotypes with the same number of cells in both compartments were excluded from analysis. Clonotypes with cells in one organ only were termed exclusive; those with more cells in one compartment than the other were termed biased.

### Statistics

R package “ggpubr” was used to generate pairwise comparisons with settings “method = ‘wilcoxon’, p.adjust.method = ‘holm’” for Mann-Whitney U tests with Holm correction and “method = ‘signs’, p.adjust.method = ‘holm’” for paired Wilcoxon signed-rank tests with Holm correction. Correlations and p values were calculated by Pearson’s product-moment using base R cor.test on clonotype count data prior to log-transformation.

## Data Availability

All data produced in the present study are available upon reasonable request to the authors.

## Acknowledgements

We thank the patients who participated in the study. We thank the Mount Sinai Human Immune Monitoring Center for 10x single cell library preparation and the flow cytometry core for support. This work was supported by NIH/NIDDK grants R01 DK112978 (J.J.F) and R01 DK123749 (S.M). This work was supported in part through the computational and data resources and staff expertise provided by Scientific Computing and Data at the Icahn School of Medicine at Mount Sinai and supported by the Clinical and Translational Science Awards (CTSA) grant UL1TR004419 from the National Center for Advancing Translational Sciences. Research reported in this publication was also supported by the Office of Research Infrastructure of the National Institutes of Health under award number S10OD026880 and S10OD030463. The content is solely the responsibility of the authors and does not necessarily represent the official views of the National Institutes of Health.

---

**Supplemental Figure 1.**
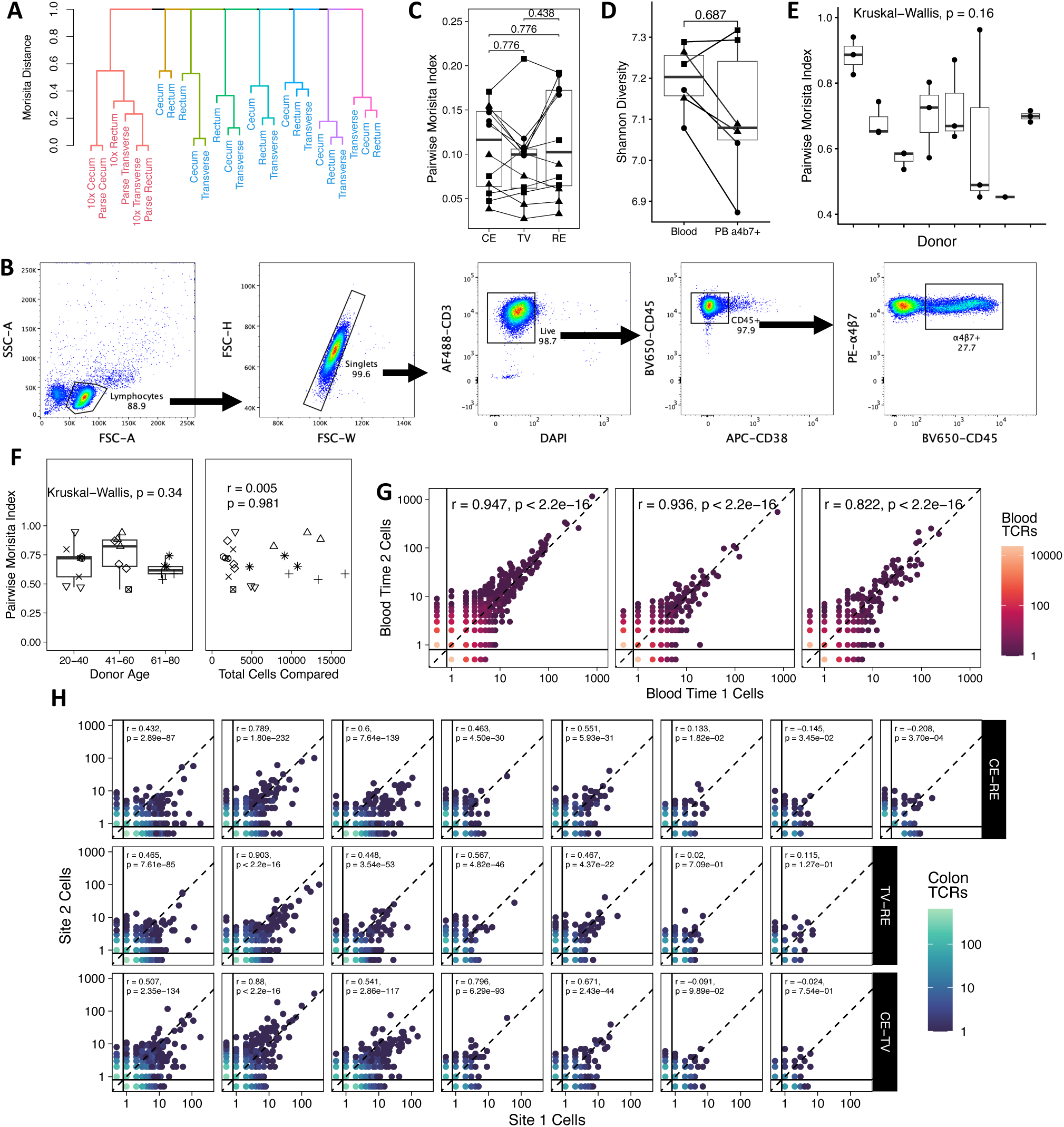
TCR Repertoire Sharing is Consistent Across Methods, Unrelated to Age or Sample Size, and Conserved Across Time Points and Colon Sites. (A) Hierarchical clustering of Morisita distance groups same donor samples together across scTCR methods. Samples in red had T cells from each site split and sequenced according to indicated method. Samples in blue were sequenced by one method only. (B) Gating strategy for isolation of integrin α4β7+ T cells from peripheral blood for single cell profiling. (C) Blood-colon repertoire similarity does not differ between cecum, transverse colon, and rectum in healthy controls (n=3). Each point is a pairwise repertoire comparison between colon and blood. Each colon site produced 4 comparisons to autologous blood samples across 2 enrichment statuses and 2 timepoints. Lines connect colon sites from the same donor that are compared to the same blood sample. p value by Wilcoxon signed rank test and corrected for multiple comparisons by Holm method. (D) Blood fractions do not differ in TCR repertoire Shannon diversity. All samples (n=3 donors) rarefied to lowest sample size. Each point represents one sample repertoire, shape indicates donor. Lines connect paired blood fractions from same donor and timepoint. p value by Wilcoxon signed rank test. (E) Colon site-site sharing does not significantly vary among donors. p value calculated by Kruskal-Wallis test. (F) Clonal sharing between colon sites is not significantly associated with donor age or sample size. Each point is a comparison between two repertoires, shape represents donor. Age p value calculated by Kruskal-Wallis. Sample size correlation and p values calculated by Pearson’s product-moment. (G) Blood clonotypes have similar expansion across time points within each donor. Point location indicates number of cells detected at each site per clonotype, point color indicates number of clonotypes with that cell distribution. Clonotypes with 0 cells in one compartment represented as 0.5 (below solid lines) for visualization on log-scale axes, dashed lines indicate equal clone size in both sites. p value and correlation calculated by Pearson’s product-moment for each comparison. (H) Colon clonotypes are shared across anatomic sites within each donor, plots as in (D). p value and correlation calculated by Pearson’s product-moment for each comparison. Abbreviations: CE = cecum, TV = transverse colon, RE = rectum.

**Supplemental Figure 2.**
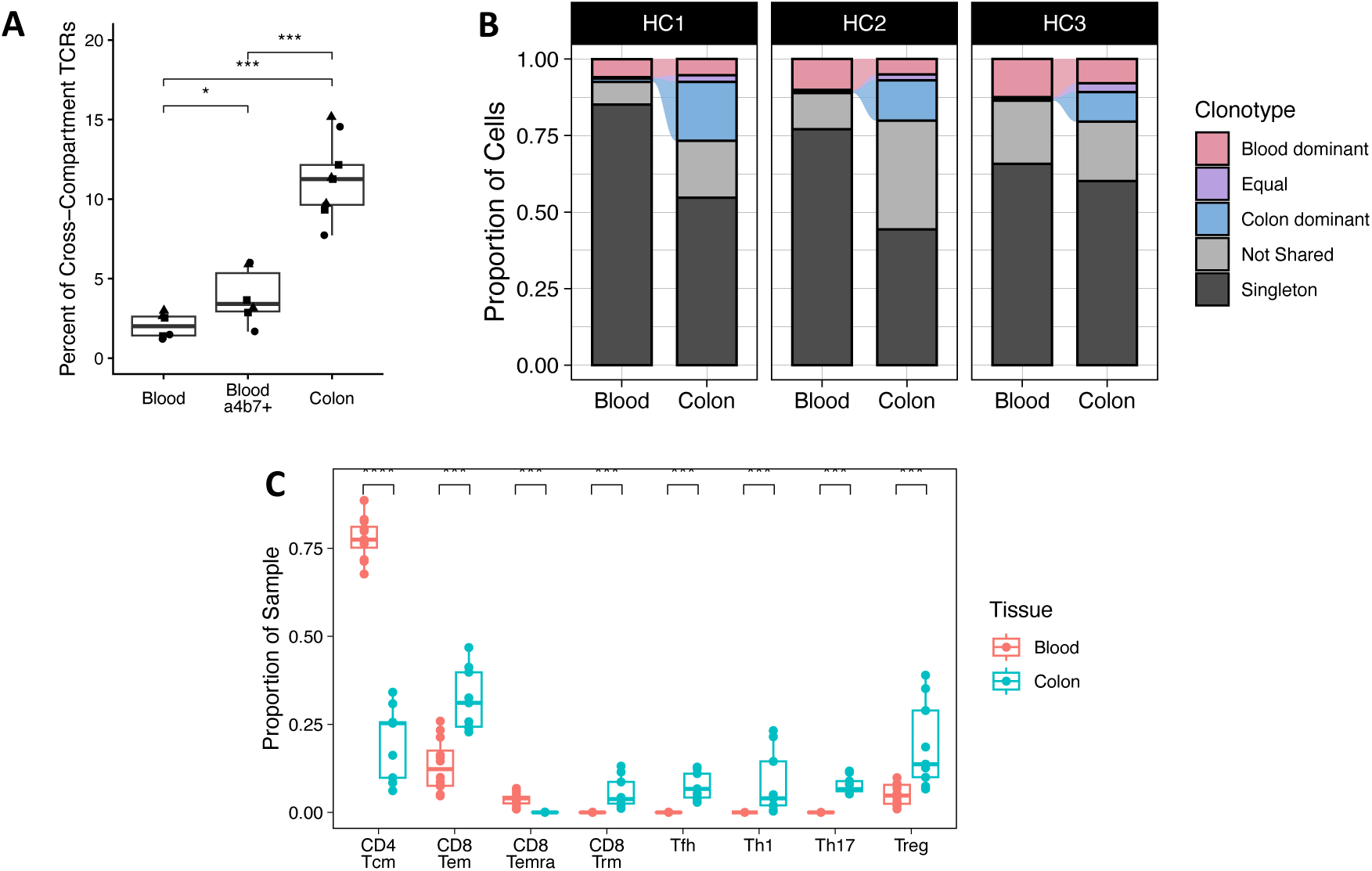
Blood-Colon TCR Sharing is Greater in Colon than in Blood. (A) Colon repertoires are made up of a greater share of cross-compartment shared clonotypes. Each point represents one repertoire, shape indicates donor (n = 3). p values for pairwise comparisons calculated by Mann-Whitney U test and corrected for multiple comparisons by Holm method. *p < 0.05, ***p < 0.001. (B) Cells of shared clonotypes make up a greater share of colon than blood. Stacked bar chart of proportion of cells in each compartment colored by clonotype sharing across compartments. Dominance assigned to the compartment with greater share of cells for each clonotype. Clonotypes that are expanded within one compartment but not shared are indicated as “Not Shared”, clonotypes with only one cell are termed “Singleton”. (C) Proportion of each phenotype per sample differs between colon and blood samples. p values for calculated by Mann-Whitney U test and corrected for multiple comparisons by Holm method. ***p < 0.001, ****p < 0.0001

**Supplemental Figure 3.**
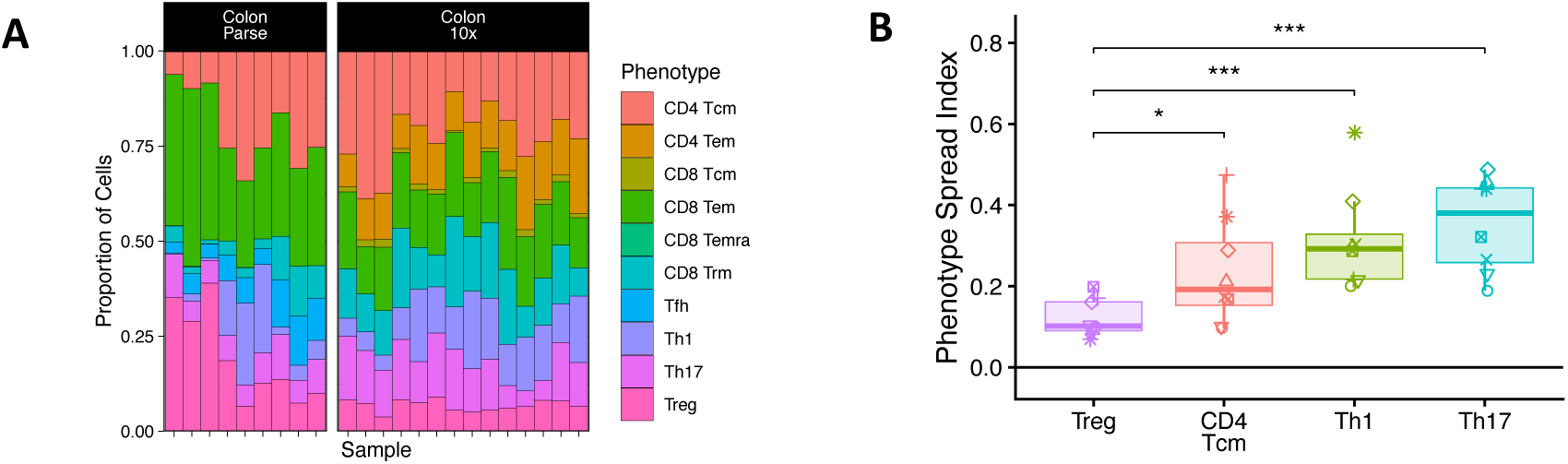
Colon T Cell Phenotypes Are Simlar Across Methods, Treg Clonal Groups Show Lower Phenotype Variation. (A) T cell phenotype for colon samples processed by split-pool (Parse Biosciences) and flow-cell (10x) barcoding. (B) Tregs have lower clonotype phenotype breadth across colon than Th17. Phenotype spread index indicates evenness of clonal T cell spread across phenotypes from all samples within a donor. p values calculated by Mann-Whitney U test and corrected for multiple comparisons by Holm method. *p < 0.05, ***p < 0.001

**Supplemental Figure 4.**
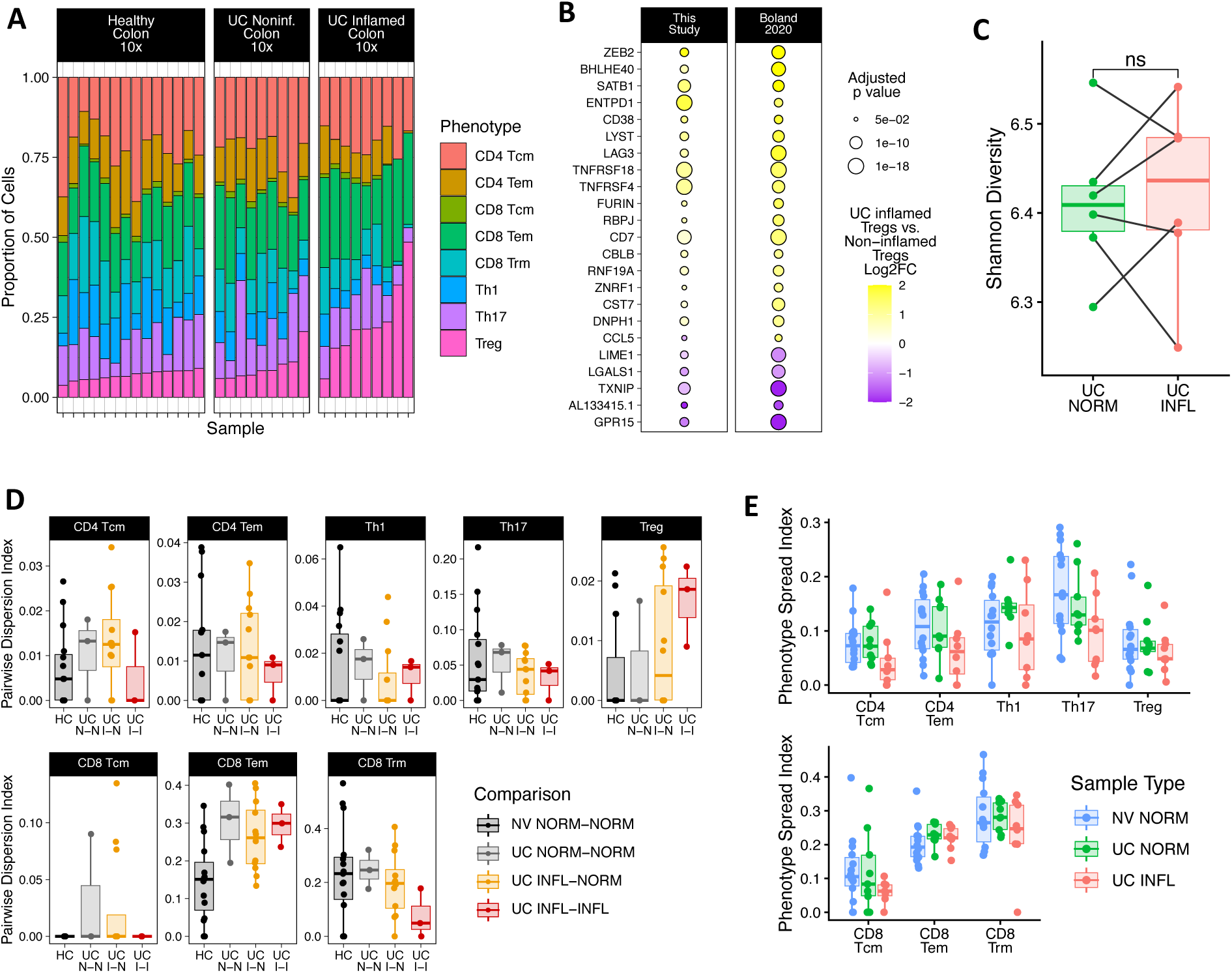
UC Colon Inflammation Alters Dispersion Trends of Treg and CD8 Trm Clonal Groups and Phenotype Spread of CD4 Clonal Groups. (A) T cell phenotype for healthy (n = 5 donor) and UC (n = 6 donor) colon samples processed by 10x. (B) Shared transcriptomic signature of UC inflamed colon Tregs vs. healthy uninflamed Tregs. All genes listed in both datasets included. Included genes meet significance cutoff of FDR = 0.05. (C) Paired TCR diversity comparison by Shannon entropy. All samples with matching inflammation status were combined for each donor, then downsampled to the smallest sample size. Lines connect samples from same donor. p value calculated by Wilcoxon signed-rank test. (D) Tregs and CD8 Trm show altered pairwise migration index in inflamed colon vs uninflamed. Pairwise dispersion index indicates evenness of clonal T cell spread across site pairs with matching phenotype labels. (E) Phenotype spread index is lower in CD4 phenotypes in UC inflamed colon compared to healthy donors.

**Table Sl.**
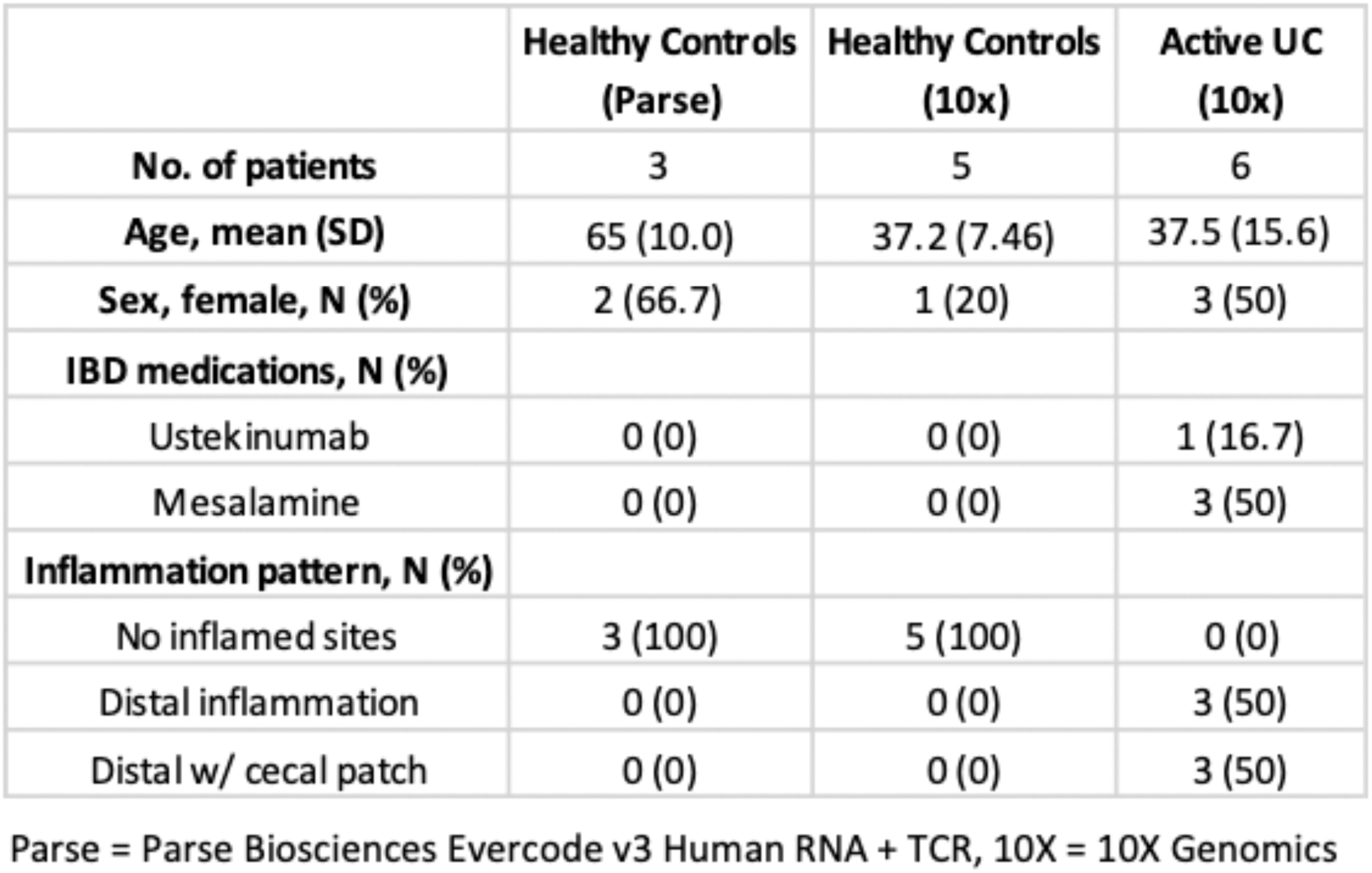
Donor Characteristics.

